# Real-world, Prospective, and Multicenter Validation of a microRNA-based Thyroid Molecular Classifier

**DOI:** 10.1101/2020.10.27.20217356

**Authors:** Marcos Tadeu dos Santos, Bruna Moretto Rodrigues, Satye Shizukuda, David Livingstone Alves Figueiredo, Giulianno Molina de Melo, Rubens Adão da Silva, Claudio Fainstein, Gerson Felisbino dos Reis, Rossana Corbo, Helton Estrela Ramos, Fernanda Vaisman, Mário Vaisman

## Abstract

**Background:** The diagnosis of cancer in thyroid nodules with indeterminate cytology (Bethesda III/IV) is challenging as fine-needle aspiration (FNA), the gold standard method, has limitations, and these cases usually require diagnostic surgery. As approximately 77% of these nodules are not malignant, a diagnostic test accurately identifying benign thyroid nodules can reduce surgery rates. We have previously reported the development and validation of a microRNA-based thyroid molecular classifier for precision endocrinology (mir-THYpe) with high sensitivity and specificity, which could be performed directly from readily available cytological smear slides without the need for a new dedicated FNA. We sought to evaluate whether the use of this test in real-world clinical routine can reduce the rates of surgeries for Bethesda III/IV thyroid nodules and analyze the test performance.

**Methods:** We designed a real-world, prospective, multicenter cohort study. Molecular tests were performed in a real-world clinical routine with samples (FNA smear slides) prepared at 128 cytopathology laboratories. Patients were followed-up from March 2018 until surgery or until March 2020 (for those patients not recommended for surgery). The final diagnosis of thyroid tissue samples was retrieved from postsurgical anatomopathological reports.

**Results:** After applying the exclusion criteria, 435 patients (440 nodules) classified as Bethesda III/IV were followed-up. The rate of avoided surgeries was 52.5% for all surgeries and 74.6% for “potentially unnecessary” surgeries. After the statistical treatment of non-resected test-negative samples, the test achieved 89.3% sensitivity (95% CI 82–94.3), 81.65% specificity (95% CI 76.6–86), 66.2% positive predictive value (95% CI 60.3–71.7), and 95% negative predictive value (95% CI 91.7–97) at 28.7% (95% CI 24.3–33.5) cancer prevalence. The test influenced 92.3% of clinical decisions.

**Conclusions:** The reported data demonstrate that the use of the microRNA-based classifier in the real-world can reduce the rate of thyroid surgery with robust performance and significantly influence clinical decision-making.

## INTRODUCTION

Thyroid cancer has increased in the last few decades, and it ranks as the ninth most-incident cancer worldwide.^1^ Although there is consensus that overdiagnosis is the main reason for increased incidence ^2,3^ mainly due to incidental thyroid nodules found on imaging screenings performed for reasons other than thyroid disease evaluation,^4^ there has also been a true increase in the occurrence of thyroid cancer.^5^

Fine-needle aspiration (FNA) cytology is the current gold standard for triaging patients with suspicious thyroid nodules detected clinically or on ultrasound (US),^6-8^ and the six-tier Bethesda System for Reporting Thyroid Cytopathology attempts to standardize cytopathologic analysis.^9^

Approximately 64% of histologically diagnosed thyroid cancer nodules were initially classified on FNA cytology as Bethesda V or VI, with a combined risk of malignancy (RoM) of 91% (only 1.1 surgeries required to detect one case of cancer). Although the proportion of histologically diagnosed thyroid cancer nodules initially classified on FNA cytology as “indeterminate” (Bethesda III and IV) is lower (∼29%), the combined RoM for these two classes is about 23% only,^10^ implying that around 77% of surgeries are potentially unnecessary and could have been reconsidered or avoided (4.4 surgeries required to detect one case of cancer).

Molecular testing has emerged as an option to reduce the need for diagnostic surgery in indeterminate thyroid nodules,^8^ recommended by the American Thyroid Association (ATA)^11^ and National Comprehensive Cancer Network (NCCN)^12^ guidelines. We have recently reported the development and validation of a new microRNA-based thyroid molecular classifier test for a precision endocrinology (mir-THYpe) with high sensitivity and specificity, which could be performed directly from readily available cytological smear slides at a significantly lower cost without the need for a new dedicated FNA.^13,14^

To evaluate the impact of mir-THYpe results on reducing the rates for surgery in the real-world clinical settings and analyze its performance on Bethesda III and IV nodules, we present data of a real-world, prospective, and multicenter study.

## METHODS

### Study Population and Design

As we conducted a real-world evidence study, there was no specific method of patient recruitment. Patients eligible for this study had a signed medical prescription for the mir-THYpe test in a real-world clinical routine with at least one thyroid nodule previously biopsied by FNA and classified as Bethesda III or IV. Patients were 18 years or older, provided written informed consent, and paid out-of-pocket for the test. The study was approved by the board of the investigational ethics committee.

The mir-THYpe molecular test was performed, and the report was released. Regardless of the results (positive or negative for malignancy), patients were assigned to one of the two groups according to the physician’s recommendation: Group A underwent a thyroidectomy (tests performed March 2018–March 2020); Group B avoided thyroid surgery (tests performed March 2018–October 2019, follow-up in March 2020). Patients who initially received no recommendations for surgery but later underwent thyroidectomy (regardless of the reason) were included in Group A.

Patients who underwent thyroid surgery but for whom we could not obtain original postsurgical anatomopathological (AP) reports were excluded. Samples in which the FNA cytology report and other characteristics, such as size and location, did not correlate with the nodule described in the AP report were also excluded from the performance analysis.

### Samples

The FNA cytology Bethesda classes and the postsurgical histological classification of resected nodules were not revised by other pathologists. The FNA smear slides of the 440 samples (used for the mir-THYpe test – Figure 1, box c) were prepared, and the Bethesda classes were assigned, by 128 cytopathology laboratories across 54 cities in all five regions and the Federal District of Brazil (Figure S1-Supplementary Material). All fixation and staining protocols of the FNA cytology smear slides were accepted. The stored samples were sent without temperature control (room-temperature). The 168 postsurgical AP reports with the histological classification of nodules (Figure 1, boxes h and o) were assigned by 53 pathology laboratories across 21 cities in three regions of Brazil.

**Figure 1.**
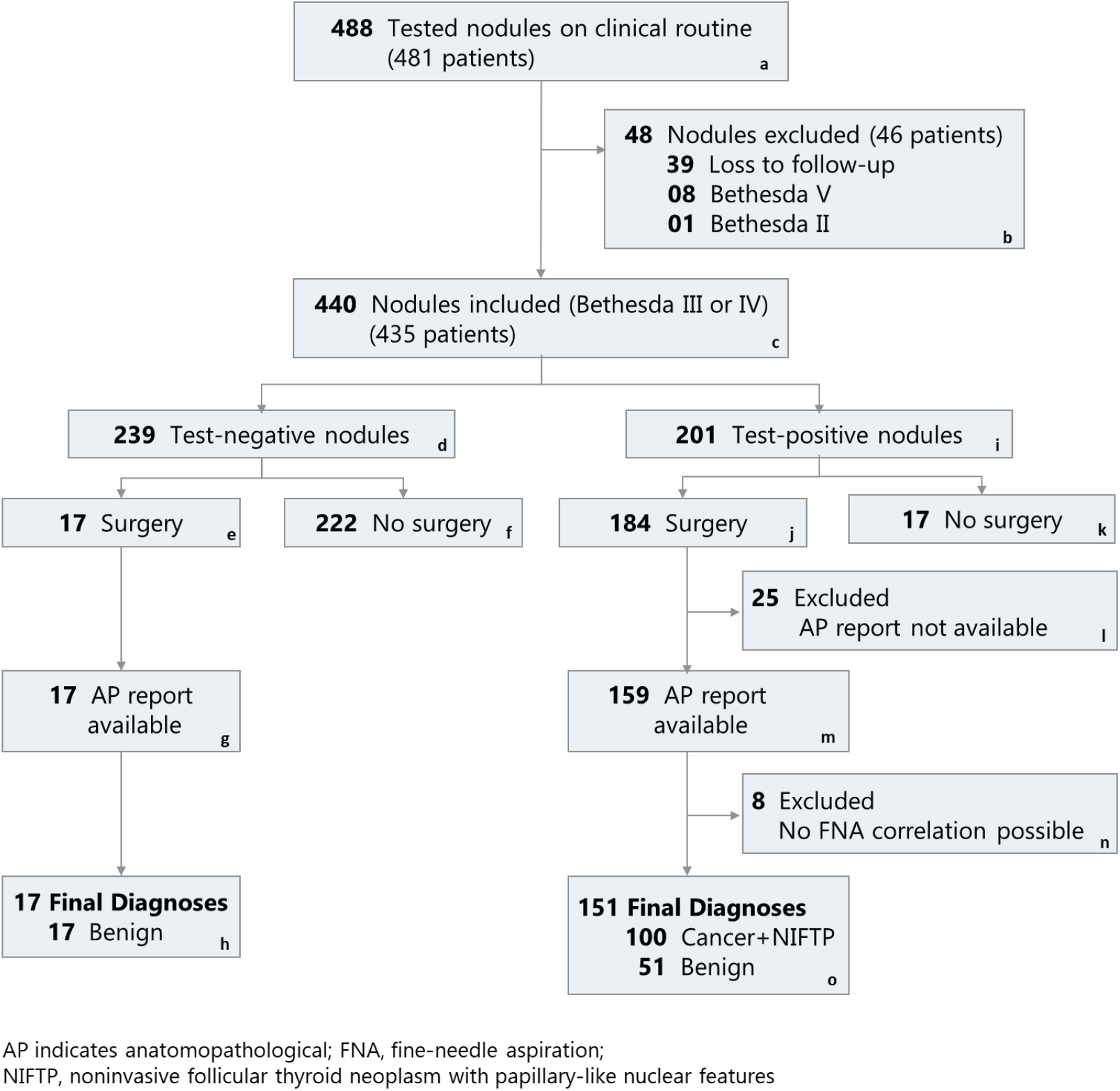
Patients and Nodules Exclusion Flow chart.

Although the indolent behavior of the noninvasive follicular thyroid neoplasms with papillary-like nuclear features (NIFTPs), its diagnosis is established only after applying stringent histological criteria.^15^ As NIFTPs require surgical treatment, positive mir-THYpe tests were considered true-positives. Data on nodule size were retrieved from postsurgical AP reports.

### Molecular Analysis

The mir-THYpe molecular classifier test was performed according to the previously described method.^13^ In brief, thyroid cells from the FNA smear slides were microdissected and submitted to manual RNA extraction, followed by reverse-transcription cDNA synthesis, preamplification stage, and real-time qPCR. The normalized Ct values for the expression of 11 microRNAs were submitted to a proprietary tree-based model that classified samples as “negative” or “positive” for malignancy. A 12^th^ microRNA (miR-375) was also analyzed to screen for medullary thyroid cancer.^16^ No mutation or DNA analyses were performed on the samples.

### Statistical Analyses

Statistical analyses were performed using R software, an open-source statistical programming environment. The confidence intervals for sensitivity, specificity, and accuracy were the “exact” Clopper– Pearson confidence intervals, and the confidence intervals for the predictive values were the standard logit confidence intervals.^17^ Confidence intervals for the means were calculated using the method described by Walline.^18^

## RESULTS

### Patient and Nodule Characteristics

A total of 481 real-world patients with 488 nodules (1.01 nodule/patient) and a medical prescription for the mir-THYpe test provided written informed consent. Of these, 39 nodules (8%) were excluded due to loss to follow-up. Eight nodules (1.6%) classified as Bethesda V and one nodule (0.2%) classified as Bethesda II were also excluded (Figure 1, box b). The remaining 440 nodules (from 435 patients; 1.01 nodules/patient) classified as Bethesda III and IV, fulfilled all initial inclusion criteria and were categorized according to the mir-THYpe test results (Figure 1 and Table 1).

**Table 1.** Demographic and Clinical characteristics of the study cohort.

| Variable | Total | mir-THYpe test result |  |
| --- | --- | --- | --- |
|  |  | Negative | Positive |
| Cohort, No. (%) |  |  |  |
| Nodules Bethesda III/IV | 440 | 239 (54.3) | 201 (45.7) |
| Unique Patients<sup>a</sup> | 435 | - | - |
| Sex, No. (%) |  |  |  |
| Male | 79 (18) | 50 (63.3) | 29 (36.7) |
| Female | 361 (82) | 189 (52.4) | 172 (47.6) |
| Age, No. (%), y |  |  |  |
| 20-54 | 252 (57.3) | 124 (49.2) | 128 (50.8) |
| ≥ 55-90 | 188 (42.7) | 115 (61.2) | 73 (38.8) |
| Bethesda Classes<sup>b</sup>, No. (%) |  |  |  |
| AUS/FLUS (III) | 247 (56.1) | 126 (51) | 121 (49) |
| FN/SFN (IV) | 193 (43.9) | 113 (58.5) | 80 (41.5) |
| Nodule size<sup>c</sup>, No. (%), cm |  |  |  |
| < 2.00 | 145 (86.3) | 12 (70.6) | 133 (88.1) |
| 2.00-4.00 | 21 (12.5) | 4 (23.5) | 17 (11.3) |
| > 4.00 | 2 (1.2) | 1 (5.9) | 1 (0.7) |
| Nodule Lobe Position, No. (%) |  |  |  |
| Left | 175 (39.8) | 83 (47.4) | 92 (52.6) |
| Right | 236 (53.6) | 139 (58.9) | 97 (41.1) |
| Isthmus | 29 (6.6) | 17 (58.6) | 12 (41.4) |

<sup>a</sup> Five patients with 2 nodules. One patient with both nodules with negative result. One patient with both nodules with positive result. Three patients with one nodule with negative and the other with a positive result.
<sup>b</sup> According to the origin pathology lab (real-world).
<sup>c</sup> Nodule sizes were retrieved only by the postsurgical AP reports.
AUS/FLUS, atypia of undetermined significance/follicular lesion of undetermined significance; FN/SFN, follicular or oncocytic (Hürthle cell) neoplasm/suspicious for a follicular or oncocytic (Hürthle cell) neoplasm; No, number of

Although the mir-THYpe test was negative for malignancy in 239 samples (Benign Call Rate [BCR] of 54.3%), 17 nodules (7.1%) underwent thyroidectomy (Group A), (9 previously classified as Bethesda III [52.9%] and 8 as Bethesda IV [47.1%]). The mean age of these 17 patients was 49 years, the mean nodule size was 1.8 cm, and the average test-to-surgery time was 170 days; for details, see Table S1 in the Supplementary Material. The mean age of the other 222 patients (92.9%) (Group B) was 52 years. They received no medical recommendation for surgery after the mir-THYpe test result (as of March 2020); the mean follow-up time was 412.9 days. Of these, 117 nodules (52.7%) were classified as Bethesda III and 105 (47.3%) as Bethesda IV; Table S2 in the Supplementary Material presents the 95% confidence intervals and ranges. Test-to-surgery and follow-up times are described in Table S3 in the Supplementary Material.

The mir-THYpe test was positive for malignancy in 201 samples (Positive Call Rate [PCR] of 45.7%), and 184 nodules (91.6%) were surgically treated (Group A). However, 25 nodules (13.6%) were excluded due to lack of access to the original postsurgical AP report, and 8 samples (4.3%) were excluded because the punctured nodule described in the FNA cytology did not correlate with the nodule(s) described in the postsurgical AP report. Of the remaining 151 resected nodules (82.1%), 83 were previously classified as Bethesda III (55%) and 68 as Bethesda IV (45%). The mean age of the patients was 49 years, the mean nodule size was 1.13 cm, and the average test-to-surgery time was 79 days. The average age of the other 17 patients (8.4%) (Group B) was 54 years, and the patients were not surgically treated (as of March 2020), despite a positive result; the mean follow-up time was 359.6 days. Among them, 13 nodules (76.5%) were classified as Bethesda III and 4 (23.5%) as Bethesda IV; Table S2 in the Supplementary Material presents the 95% confidence intervals and ranges. Test-to-surgery and follow-up times are described with periods in Table S3 in the Supplementary Material.

Overall, the “typical” profile of a real-world patient referred for the mir-THYpe test was female (82%), younger than 55 years (57.3%) with a Bethesda III (56.1%) thyroid nodule <2 cm (86.3%). Table 1 displays the demographic and clinical characteristics of the study cohort.

### Test Performance

Table 2 summarizes the test performance across all 168 resected nodules according to the histopathological type found post-surgery.

**Table 2.** Performance of microRNA-based molecular classifier according to histopathological subtype.

| mir-THYpe® result / Postsurgical tissue | FNA smear slides Bethesda classes |  |  |
| --- | --- | --- | --- |
|  | AUS/FLUS (III) | FN/SFN (IV) | Total |
| **Test-Negative nodules** | **9** | **8** | **17** |
| **Malignant** - False Negatives, No (%) | **0 (0%)** | **0 (0%)** | **0 (0%)** |
| **Benign** - True Negatives, No (%) | **9 (100%)** | **8 (100%)** | **17 (100%)** |
| Hürthle cell adenoma | 2 | 2 | 4 |
| Follicular Adenoma | 1 | 3 | 4 |
| Adenomatous goiter / follicular hyperplasia | -- | 2 | 2 |
| Colloid goiter | 5 | 1 | 6 |
| Hashimoto's thyroiditis | 1 | -- | 1 |
| **Test-Positive nodules** | **83** | **68** | **151** |
| **Malignant** - True Positives, No (%) | **52 (62.7)** | **48 (70.6)** | **100 (66.2%)** |
| Papillary carcinoma, usual type | 23 | 7 | 30 |
| Papillary carcinoma, follicular variant | 16 | 30 | 46 |
| Papillary carcinoma, Hürthle cell variant | 2 | 1 | 3 |
| NIFTP<sup>a</sup> | 6 | 3 | 9 |
| Follicular carcinoma, microinvasive | 4 | 5 | 9 |
| Follicular carcinoma, widely invasive | -- | 1 | 1 |
| Follicular carcinoma, Hürthle cell variant | -- | 1 | 1 |
| Medullary carcinoma | 1 | -- | 1 |
| **Benign** - False Positives, No (%) | **31 (37.3)** | **20 (29.4)** | **51 (33.8%)** |
| Hürthle cell adenoma | 4 | 3 | 7 |
| Follicular Adenoma | 10 | 11 | 21 |
| Adenomatous goiter / follicular hyperplasia | 9 | 5 | 14 |
| Colloid goiter | 6 | 1 | 7 |
| Hashimoto's thyroiditis | 2 | -- | 2 |
| **Total** | **92 (54.8%)** | **76 (45.2%)** | **168 (100%)** |

<sup>a</sup> Considering positive test result for NIFTP as correct classification.
NIFTP, Noninvasive follicular thyroid neoplasm with papillary-like nuclear features.
AUS/FLUS, atypia of undetermined significance/follicular lesion of undetermined significance; FN/SFN, follicular or oncocytic (Hürthle cell) neoplasm/suspicious for a follicular or oncocytic (Hürthle cell) neoplasm; No, number of.

All 17 samples with negative results treated with surgery, regardless of Bethesda class, were confirmed as benign lesions (true-negatives) in the postsurgical AP reports, achieving an overall Negative Predictive Value (NPV) of 100% (one-side 97.5% CI 80.5–100). This result must be interpreted with caution because the sample size was too small (patients with a negative test usually do not receive a surgery recommendation). This result included four Hürthle cell adenomas (23.5% [95% CI 6.8–49.9]).

Among the 151 resected nodules with positive results included in the final analysis, 100 nodules were confirmed as cancer/NIFTP lesions (true-positives) in the postsurgical AP report, including nine NIFTPs (9% [95% CI 4.2–16.4]) and one medullary carcinoma (1% [95% CI 0.02–5.5], achieving an overall Positive Predictive Value (PPV) of 66.2% (95% CI 60.3–71.7). Splitting the results by cytological class when the mir-THYpe test was positive (Table 3), 52 of the 83 Bethesda III (62.7%), and 48 of the 68 Bethesda IV nodules (70.6%) were accurately classified. Fifty-one nodules showed false-positive results due to benign tissue identified in the postsurgical AP reports (33.8% [95% CI 26.3–41.9]).

**Table 3.** True-positive performance of commercial molecular tests on prospective studies and FNA alone performance on Bethesda III and IV nodules.

| *Bethesda Class* | Samples with test-positive result, No<br>(True Cancer/NIFTP histology, No) |  |  | True-positive rate, % (95% CI) |  |  | Study Type | Sample Type |
| --- | --- | --- | --- | --- | --- | --- | --- | --- |
|  | AUS/FLUS (III) | FN/SFN (IV) | III and IV | AUS/FLUS (III) | FN/SFN (IV) | III and IV |  |  |
| **mir-THYpe** | 83 (52) | 68 (48) | 151 (100) | 62.7 (51.3-73) | 70.6 (58.3-81) | 66.2 (58.1-73.7) | Real-World | FNA Smear Slide |
| **ThyroSeq v3<sup>19</sup>** | 50 (32) | 47 (32) | 97 (64) | 64 (49.2-77.1) | 68.1 (52.9-80.9) | 66 (55.6-75.3) | Trial | New FNA |
| **Afirma GSC<sup>20</sup>** | 114 (51) | 76 (36) | 190 (87) | 44.7 (35.4-54.3) | 47.4 (35.8-59.1) | 45.8 (35.5-53.1) | Trial | New FNA |
| **FNA alone<sup>10</sup>** | 957 (152) | 1791 (468) | 2748 (620) | 15.9 (13.6-18.3) | 26.1 (24.1-28.2) | 22.6 (21-24.2) | Meta-analysis | FNA Smear Slide |

CI, Confidence Interval; FNA, fine-needle aspiration
AUS/FLUS, atypia of undetermined significance/follicular lesion of undetermined significance; FN/SFN, follicular or oncocytic (Hürthle cell) neoplasm/suspicious for a follicular or oncocytic (Hürthle cell) neoplasm; No, number of

Overall, the test accuracy was 70% (95% CI 62–76.5) as it accurately classified 117 of the 168 resected samples, regardless of test results.

Although expected, the small number of resected test-negative nodules impaired the realistic calculation of the sensitivity and NPV (both at 100%) and increased the disease prevalence to unrealistic values (60%). To minimize this effect, we performed a theoretical calculation applying the sensitivity observed during the validation study^13^ (94.6%) to the 222 non-resected nodules with test-negative results (Figure 1, box f), resulting in 210 true-negatives and 12 false-negatives. In this scenario, the mir-THYpe test achieved 89.3% sensitivity (95% CI 82–94.3), 81.65% specificity (95% CI 76.6–86), 66.2% positive predictive value (PPV) (95% CI 60.3–71.7), and 95% negative predictive value (NPV) (95% CI 91.7–97) at a 28.7% (95% CI 24.3– 33.5) disease prevalence (Table 4).

**Table 4.** Calculated Performance of microRNA-based molecular classifier in cytologically indeterminate thyroid nodules.

| *Result* | Postsurgical tissue class, No |  | Test performance |  |  |
| --- | --- | --- | --- | --- | --- |
|  | Cancer+NIFTP | Benign | Analytical parameter | % | 95% CI |
| **Positive** | 100 | 51 | **Sensitivity** | 89.3 | 82-94.3 |
|  |  |  | **Specificity** | 81.6 | 76.6-86 |
| **Negative** | 12<sup>a</sup> | 210<sup>a</sup> +17 (227)<sup>a</sup> | **NPV** | 95.0 | 91.7-97 |
|  |  |  | **PPV** | 66.2 | 60.3-71.7 |

<sup>a</sup>Theoretical values, considering the published sensitivity<sup>13</sup> of 94.6% for the 222 test-negative not surgically resected nodules.
CI, Confidence Interval; NPV/PPV, Negative/Positive Predictive Value; NIFTP, Noninvasive follicular thyroid neoplasm with papillary-like nuclear features; No, number

### Comparison with other Thyroid Molecular Classifier Tests

To evaluate the real-world utility of the mir-THYpe test, we performed a comparison with the results of prospective studies on the latest version of two leading commercially available thyroid molecular classifier tests: ThyroSeq v3^19^ and Afirma GSC.^20^ We also compared the molecular test results with the performance of the FNA alone using data from a meta-analysis^10^ (Table 3).

The true-positive rate of the mir-THYpe test (66.2%) for Bethesda III and IV cytology was very similar to that in the ThyroSeq v3 prospective study (66%), but the number of tested samples in the present study was 35% higher (151 vs. 97). The mir-THYpe showed significantly better performance than the results for Afirma GSC (45.8%), and all molecular tests showed better results than those obtained with FNA cytology alone (22.6%). It is noteworthy that among the compared molecular classifiers, ours is the only study to perform the test exclusively with FNA smear slides and a real-world design.

### Influence on Clinical Decisions and Surgery Reduction Rates

The mir-THYpe test influenced 93% of clinical decisions when the result was negative for malignancy (222 nodules with no medical recommendation for surgery, out of 239 test-negatives). The positive test results influenced 91.5% of clinical decisions (184 nodules treated surgically, out of 201 test-positives). Overall, the mir-THYpe test influenced 92.3% of clinical decisions (406 out of 440 cases).

To calculate the rates of total avoided surgeries among the 440 eligible patients, we assumed that 423 would have undergone thyroidectomy if no molecular test was available (we subtracted 17 patients not treated surgically, despite a positive result — Figure 1, box k). Of these, 222 patients avoided surgery (Figure 1, box f), resulting in 52.5% (95% CI 47.6–57.3) of all surgeries.

Following Figure 1, we considered that 280 patients (17 [box e] + 222 [box f] + 51 [box o]) had a benign nodule and would undergo a potentially unnecessary surgery if no molecular test was available. As we could not assume that all 222 samples from box f were truly benign, we applied the sensitivity observed in the validation study^13^ (94.6%) and estimated that around 210 samples were true-negative nodules in this cohort, resulting in a total of 268 patients who would have underwent a potentially unnecessary surgery if no molecular test was available. Considering that only 68 patients (17 [box e] + 51 [box o]) were surgically treated, the mir-THYpe test avoided 74.6% (95% CI 69–79.2) of potentially unnecessary surgeries.

## DISCUSSION

The present study aims to prospectively evaluate the real-world performance, impact on clinical decisions, and the true potential of a previously described microRNA-based thyroid molecular classifier for avoiding surgeries in a multicenter cohort.^13^

Although randomized controlled trials (RCTs) are considered the gold standard for analyzing the efficacy of therapies, they are limited to a subset of patients who are not fully representative of the unselected real-world patients.^21-23^ Real-world evidence studies better represent routine practice when compared with idealized conditions of RCTs, thus providing a valuable reflection of the range and distribution of patients observed in clinical practice.^22-24^ A real-world evidence study design can help physicians understand the practical use of molecular tests, providing a clear view of what to expect.

One main advantage was that the submitted specimens belonged to patients with a real clinical indication for testing and/or a desire to avoid surgery, thus avoiding the bias of including samples based on the Bethesda classification only. Moreover, bias was minimized as the patients paid out-of-pocket for the tests. Another advantage was that we could not control where the samples came from; FNA smear slides were sent at room-temperature from anywhere, and there were no boundaries to guarantee a highly heterogeneous cohort. The FNA smear slides were prepared at 128 cytopathology laboratories, ensuring a highly heterogeneous sample cohort and robustness to analyze the samples (prepared using different fixation and staining protocols by various operators). As summarized in Table 1, patient demographics and nodule characteristics were in accordance with expected real-world proportions, illustrated in the “typical” patient profile described above. Lastly, the patient and physician received the molecular test results before deciding the next step (unlike blinded studies in which all nodules are resected, and test influence is unmeasurable). Therefore, we could follow the patients and observe the real impact of their more-informed clinical decisions and test utility on surgery avoidance. The high impact of the mir-THYpe test on real-world clinical decision-making (92.3% overall) re-emphasizes that the Bethesda categories III and IV present a challenge for physicians to decide the best option for their patients. 74.5% of potentially unnecessary surgeries were avoided, suggesting that the molecular test could minimize this problem and potentially improve the efficiency of the healthcare system.

However, our study had some limitations. The multicenter design made it difficult to closely monitor patients as they were not centralized to a few institutions, and there was limited access to other clinical parameters. Ideally, non-operated patients should be followed with US at least to identify possible evolutions in the nodules suggestive of management changes. Although the physicians routinely monitored the patients, we had restricted access. Therefore, we had to limit the follow-up monitoring according to whether the patient underwent thyroid surgery or not. Moreover, we did not review the patients’ FNA or AP slides in order to guarantee the real-world approach, even though we assumed the risk of potential misclassifications at the pathology laboratories. Finally, as the majority of patients who tested negative were not submitted to thyroid surgery (222/239, 93%), we did not have the statistical power to evaluate the performance of negative results (mainly the sensitivity and the NPV). Although all 17 resected nodules with negative results were confirmed benign, the sample size impaired the confidence on 100% sensitivity and NPV. However, the statistical adjustment applied (described above on Results) for the 222 test-negative samples to minimize this effect and calculate the test performance was valid as the sensitivity of a diagnostic test is a parameter not influenced by disease prevalence. Thus, on this scenario, the present study achieved an NPV of 95%, which seems realistic (similar to the NPV of 95.9% observed in the previous validation study^13^).

The statistical parameters analyzing the performance of a test are crucial in determining practical applications and adoption. ThyroSeq v3 and Afirma GSC are the two main available thyroid molecular classifiers, widely adopted worldwide with proven clinical impact. Hence, we compared the mir-THYpe test performance observed in this study with prospective studies describing the performance of the latest version of these two tests.^19,20^ Based on the true-positives for each test, the mir-THYpe achieved similar performance to the ThyroSeq v3 test (62.7% vs. 64%, 70.6% vs. 68.1%, and 66.2% vs. 66% for Bethesda III, IV, and overall, respectively) and better performance than the Afirma GSC in all scenarios (62.7% vs. 44.7%, 70.6% vs. 47.4%, and 66.2% vs. 45.8% for Bethesda III, IV, and overall) (See Table 3 for details and 95% confidence intervals). In practical and quantitative terms, when either mir-THYpe or ThyroSeq v3 is used alongside FNA indeterminate cytology, around 1.5 surgeries are required to identify 1 cancer patient. If Afirma GSC is chosen, 2.18 surgeries are required to identify 1 cancer patient (1.45 more than when using mir-THYpe) and 4.43 surgeries are required with FNA alone (2.94 more than adding mir-THYpe) (See Table S4 in the Supplementary Material). Based on the NPV of each test, the mir-THYpe achieved similar performance (95%) to ThyroSeq v3 (97%) and Afirma GSC (96%). Moreover, the sample type requested to perform the test is also relevant, because if no material was stored from the first FNA (frozen), ThyroSeq v3 and Afirma GSC need a new and dedicated FNA. Therefore, patients need to attend a restricted collection site and undergo another invasive procedure. Furthermore, delicate logistics operations are needed, as the samples must be refrigerated and delivered on time. For physicians, there is no guarantee that the new puncture will have the same cellular composition as the previous sample that indicated the need for molecular testing or that the logistics chain will maintain the sample at the appropriate temperature and ensure timely delivery. Although ThyroSeq v3 is under validation for FNA smear slides, the available results are limited to only 31 samples with different performance depending on slide staining (Papanicolau 100%, Diff-Quick 65%, and overall 81%).^25^ Furthermore, FNA smear slides were not used for the prospective study validation.^19^

In summary, the reported data demonstrate over a heterogeneous and large cohort of patients that the real-world use of the mir-THYpe test can reduce the rates of surgeries for Bethesda III and IV indeterminate cytology nodules and significantly influence clinical decisions. Moreover, the real-world test performance is robust and comparable to other well-established thyroid molecular classifiers. Furthermore, it can be performed using already available cytology smear slides at a significantly lower cost. Cost-effectiveness analyses of the test’s financial impact and healthcare system savings are under development, and together with the present data, will help to guide new policies.

## Data Availability

Due to the nature of this research, to protect participants' privacy and confidentiality, raw data are not shared publicly. Relevant data supporting the findings and conclusions of this study are available within the article and its supplementary materials. Upon a justifiable request, the share of de-identified data should be approved by the board of an investigational ethics committee.

## Author Contributions

MTS designed and supervised the study. DLAF, GMM, RAS, CF, GFR, RC, HER, FV and MV contributed with data and medical expertise. BMR and SS contributed with data collection, integration, and patients’ follow-up. MTS drafted the manuscript. MTS and BMR contributed as molecular biologists. All authors read and approved the final version.

## Acknowledgments

We are grateful to Andrei Felix for laboratory support, and the SUPERA Technology and Innovation Park team for institutional support. Finally, and most importantly, the authors would like to thank all the patients who made this study possible.

## SUPPLEMENTARY MATERIAL

**Figure S1.**
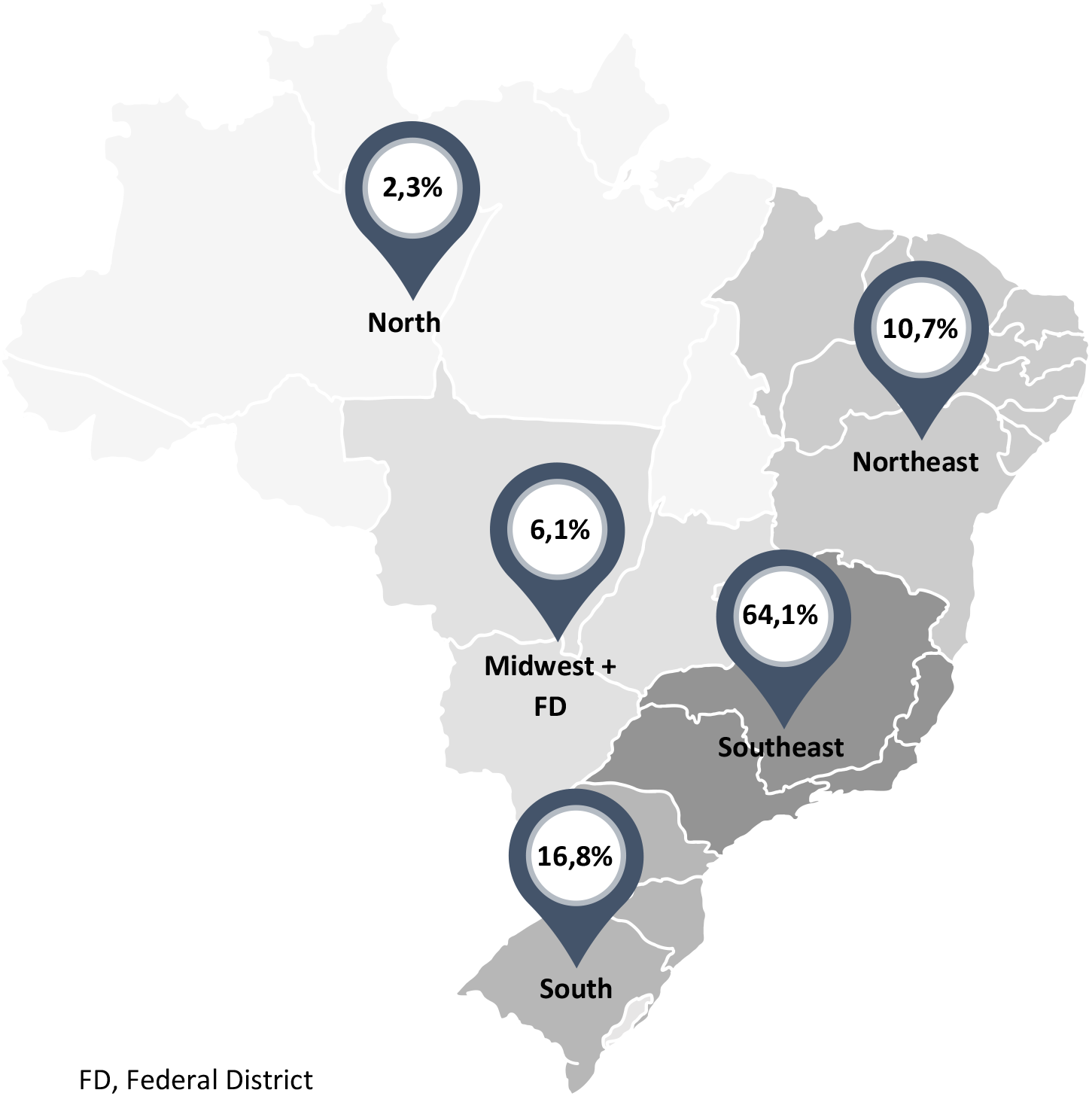
Geographic distribution of the 128 cytopathology labs where the 440 FNA smear slides tested in the study were prepared and the Bethesda categories assigned.

**Table S1.** Clinical details of the 17 patients with a test-negative result with nodules surgically resected (all true-negatives).

| Patient | Bethesda Class | Postsurgical histological subtypes | Test-to surgery (days) | Nodule size (cm) | Nodule Lobe Position | Sex |
| --- | --- | --- | --- | --- | --- | --- |
| 1 | III | Colloid goiter | 25 | 1,5 | Right | Female |
| 2 | III | Colloid goiter | 39 | 0,5 | Right | Male |
| 3 | III | Colloid goiter | 142 | 1,0 | Left | Female |
| 4 | III | Colloid goiter | 350 | 1,5 | Left | Female |
| 5 | III | Colloid goiter | 412 | 1,4 | Right | Female |
| 6 | III | Follicular Adenoma | 76 | 3,0 | Left | Female |
| 7 | III | Hashimoto's thyroiditis | 76 | 2,0 | Isthmus | Female |
| 8 | III | Hürthle cell adenoma | 78 | 5,3 | Right | Male |
| 9 | III | Hürthle cell adenoma | 182 | 3,0 | Right | Female |
| 10 | IV | Adenomatous goiter / follicular hyperplasia | 74 | 1,0 | Isthmus | Female |
| 11 | IV | Adenomatous goiter / follicular hyperplasia | 320 | 1,7 | Left | Male |
| 12 | IV | Colloid goiter | 115 | 1,5 | Isthmus | Female |
| 13 | IV | Follicular Adenoma | 52 | 1,5 | Right | Female |
| 14 | IV | Follicular Adenoma | 120 | 1,4 | Left | Female |
| 15 | IV | Follicular Adenoma | 321 | 3,0 | Left | Male |
| 16 | IV | Hürthle cell adenoma | 117 | 0,7 | Isthmus | Male |
| 17 | IV | Hürthle cell adenoma | 390 | 0,6 | Right | Female |
| *Mean* |  |  | 169,9 | 1,8 |  |  |
| *95% CI* |  |  | 101.8-238.19 | 1.19-2.41 |  |  |

CI, Confidence Intervals
Ages, range 22–70; mean 49 years (95% CI 41.1–56.8). Ages are not specified to each patient to preserve patient's confidentiality and privacy

**Table S2.** Detailed Demographic and Clinical characteristics of the study by cohort groups.

|  | mir-THYpe test result |  |
| --- | --- | --- |
|  | Negative | Positive |
| **Total** | 239 (54.3) [49.5-59] | 201 (45.7) [40.9-50.5] |
| **Group A (Surgery)** |  |  |
| Nodules resected, No (%) [95% CI] | 17 (7.1) [4.2-11.1] | 184 (91.6) [86.8-95] |
| Included, No (%) [95% CI] | 17 (100) [80.5-100<sup>a</sup>] | 151 (82.1) [75.7-87.3] |
| Bethesda III, No (%) [95% CI] | 9 (52.9) [27.8-77] | 83 (55) [46.7-63] |
| Bethesda IV, No (%) [95% CI] | 8 (47.1) [23-72.2] | 68 (45) [36.9-53.3] |
| Corrected classified, No (%) [95% CI] | 17 (100) [80.5-100<sup>a</sup>] | 100 (66.2) [58.1-73.7] |
| Age, mean (range) [95% CI] years | 49 (22-70) [41.1-56.8] | 49 (22-85) [46.8-51.2] |
| Nodule Size, mean (range) [95% CI] cm | 1.8 (0.5-5.3) [1.2-2.4] | 1.13 (0.05-4.2) [1.02-1.24] |
| Test-to-surgery, mean (range) [95% CI] days | 170 (25-412) [101.8-238.2] | 79 (16-326) [68.9-89] |
| **Group B (No-Surgery)** |  |  |
| Nodules followed-up, No (%) [95% CI] | 222 (92.9) [88.7-95.8] | 17 (8.4) [5-13.2] |
| Bethesda III, No (%) [95% CI] | 117 (52.7) [45.9-59.4] | 13 (76.5) [50.1-93.2] |
| Bethesda IV, No (%) [95% CI] | 105 (47.3) [40.6-54.1] | 4 (23.5) [6.8-49.9] |
| Age, mean (range) [95% CI] years | 52 (20-86) [50.2-53.8] | 54 (29-90) [44.7-63.2] |
| Follow-up time, mean (range) [95% CI] days | 412.9 (120-796) [388.2-437.4] | 359.6 (120-733) [266.5-452.6] |

<sup>a</sup> 97% CI, one side
CI, Confidence Intervals; No, number of

**Table S3.**
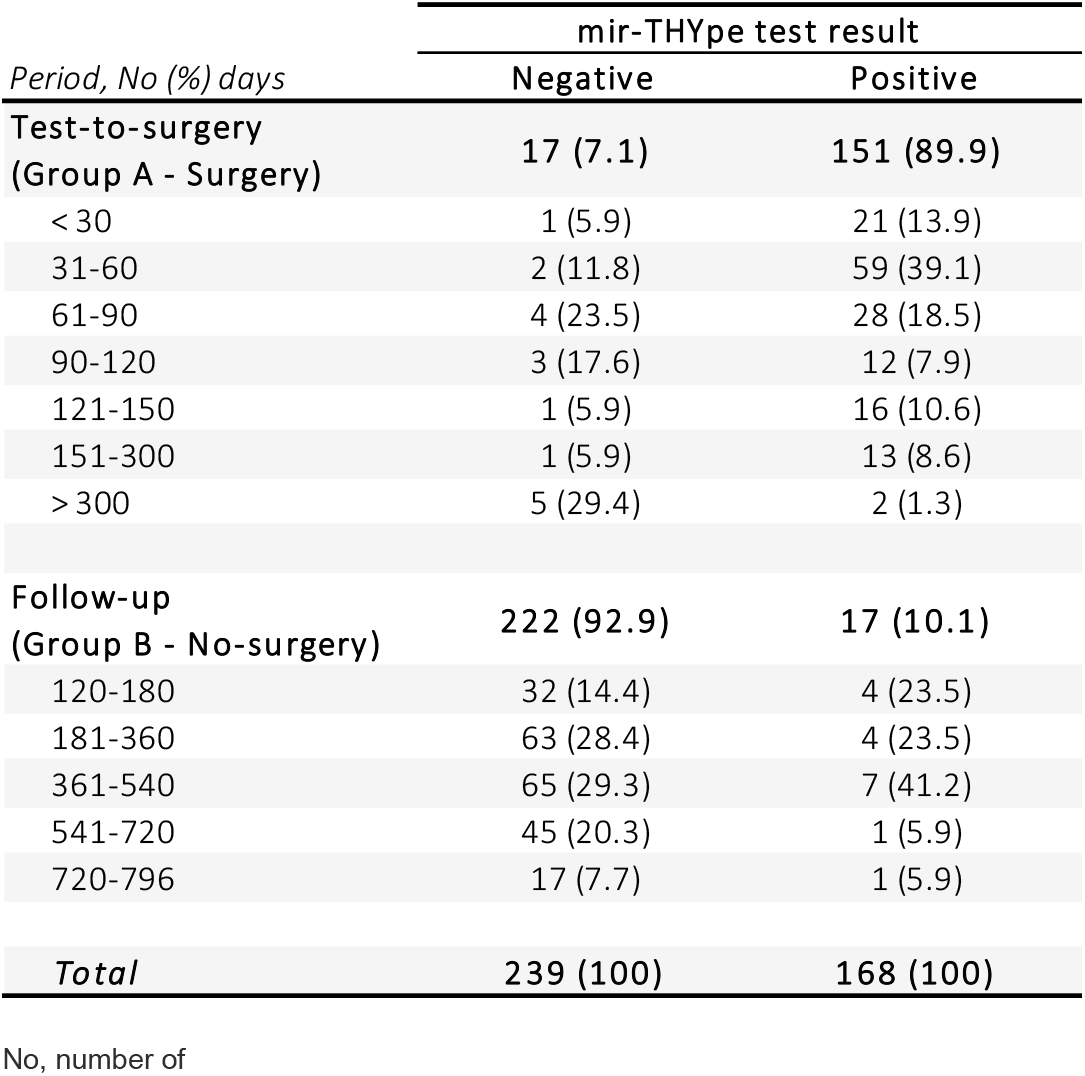
Test-to-surgery and follow-up periods of the study by cohort groups.

**Table S4.** Calculated number of surgeries needed to identify 1 case of cancer through molecular tests and FNA alone, according to the results of each study.

| *Tests / Bethesda Class* | Surgeries needed to find 1 cancer case (rates) |  |  | Differences to the mir-THYpe |
| --- | --- | --- | --- | --- |
|  | AUS/FLUS (III) | FN/SFN (IV) | III and IV |  |
| mir-THYpe | 1,60 | 1,42 | 1,51 | n.a |
| ThyroSeq v3<sup>1</sup> | 1,56 | 1,47 | 1,52 | 1,00 |
| Afirma GSC<sup>2</sup> | 2,24 | 2,11 | 2,18 | 1,45 |
| FNA alone<sup>3</sup> | 6,30 | 3,83 | 4,43 | 2,94 |

AUS/FLUS, atypia of undetermined significance/follicular lesion of undetermined significance; FN/SFN, follicular or oncocyctic (Hürthle cell) neoplasm/suspicious for a follicular or oncocyctic (Hürthle cell) neoplasm; n.a, not applicable rates, 100/% true-positive

